# Can artificial intelligence help a clinical laboratory to draw useful information from limited data sets ? Application to Mixed Connective Tissue Disease

**DOI:** 10.1101/2023.05.23.23290343

**Authors:** Daniel Bertin, Pierre Bongrand, Nathalie Bardin

**Affiliations:** Service d’Immunologie, Biogénopôle, Hôpital de la Timone, Assistance Publique-Hôpitaux de Marseille (AP-HM), Marseille, France; Aix-Marseille Univ, Laboratoire Adhésion et Inflammation, UM61, Marseille, France; INSERM U1067, France; CNRS, U7333, France; INSERM, U1076, France; Aix Marseille Univ, INSERM, INRAE, C2VN, Marseille, France

**Keywords:** machine learning, learning from data, systemic lupus erythematosus, mixed connective tissue disease, medical diagnosis, scikit-learn

## Abstract

Diagnosis is a key step of patient management. During decades, refined decision algorithms and numerical scores based on conventional statistic tools were elaborated to ensure optimal reliability. Recently, a number of machine learning tools were developed and applied to process more and more extensive data sets, including up to million of items and yielding sophisticated classification models. While this approach met with impressive efficiency in some cases, practical limitations stem from the high number of parameters that may be required by a model, resulting in increased cost and delay of decision making. Also, information relative to the specificity of local recruitment may be lost, hampering any simplification of universal models. Here, we explored the capacity of currently available artificial intelligence tools to classify patients found in a single health center on the basis of a limited number of parameters. As a model, the discrimination between systemic lupus erythematosus (SLE) and mixed connective tissue disease (MCTD) on the basis of thirteen biological parameters was studied with eight widely used classifiers. It is concluded that classification performance may be significantly improved by a knowledge-based selection of discriminating parameters.

## 1 Introduction

Diagnosis is a key step of patient management since it is required to establish prognosis and therapeutic strategy. It was soon recognized that some kind of mathematical grading could improve the reliability of conclusions and simple scoring systems were built with standard statistical methods. Well-known examples are the 5-item Apgar score that was used for decades to assess the viability of newborn babies [1] or Well’s criteria for pulmonary embolism ([2]p.19). Following the progress of medical knowledge, more and more sophisticated algorithms were developed to serve as decision-making tools. As a representative example, a set of 702 patient cases was used by a panel of experts to build an diagnostic algorithm for systemic lupus [3]. This made use of fairly refined statistical methodologies such as logistic regression and decision tree analysis. During the following years, the growing availability of computer-based tools for performing multivariate statistical analysis led to the development of elaborate classification methods making use of more and more numerous parameters and extensive data sets [4]. Thus, an artificial intelligence approach was followed to build an algorithm for the detection of atrial fibrillation on the basis of 454,789 electrocardiograms from 126,526 patients that were used to train a convolutional neural network [5]. Other authors assessed the potential of proteomic and metabolomic analysis of patients’ sera to achieve an early detection of several forms of COVID-19: after testing 894 proteins and 941 metabolites, they build a random forest machine learning model based on 29 molecules [6]. It was reported that a model built on the analysis of 2.3 million electrocardiograms could predict mortality during the next year [7]. The delimitation between standard statistical methods such as logistic regression and more recent techniques related to “artificial intelligence” or “machine learning”, such as support vector machines or neural networks [4] may be difficult to define [8], [9]. However, there is no doubt that new prediction tools share a number of properties including availability of fairly autonomous software allowing efficient processing of enormous data sets in order to yield powerful decision-making models. This evolution entails specific problems: (i) First, while it is obviously attractive to be able to combine a high number of parameters to optimize diagnosis, following this strategy may be both costly and conducive to damageable delay, if a substantial amount of time is required to determine all necessary parameters. (ii) Secondly, while it would be attractive to establish universal diagnostic rules, accounting for differences between lifestyle, local pathogens or genetic background might make site-specific models less data hungry and as efficient as universal guidelines. Indeed, as was rightly emphasized ([2]p.16) *All guidelines recognize that “one size fits all” recommendations may not apply to individual patients*. Algorithms derived from the study of a particular population may not apply to other groups [10]. (iii) Thirdly, the efficiency and facility of use of available software is usually due to the existence of control procedures such as “regularization” depending on hidden parameters. A drawback of the use of too complex models is that the significance of predictions is often difficult to assess. In other words, they may appear as “black boxes” the conclusions of which may be difficult to translate into some general wisdom [11]. Indeed, if a conclusion is drawn from a high number of correlated numbers, is may difficult to identify independent basic parameters. (iv) In line with above remarks, machine learning was reported to need more data than conventional statistical tools to achieve a comparable accuracy [12]. (v) While algorithms based on machine learning were sometimes claimed to achieve an accuracy matching human expertise, some progress remains possible, and it was suggested that human expertise could improve the efficiency of artificial intelligence [13] [14].

The purpose of the this report was to explore the potential and limitation of currently available machine learning algorithms to yield useful information by processing parameters easily accessible to a clinical immunology laboratory under realistic conditions.

As a model, we chose the differential diagnosis between systemic lupus erythematosus (SLE) and mixed connective tissue disease (MCTD). SLE is a systemic auto-immune disease with a prevalence on the order of 1/1,000 women in the USA [2]. Diagnosis is based on a set of clinical and biological criteria following regularly updated algorithms [3]. Important biological markers include anti-nuclear antibodies as revealed with immunofluorescence, anti-double stranded DNA and a number of more or less precisely defined auto-antibodies directed against a number of nuclear antigens [15]. MCTD was described as a new connective tissue disease displaying overlapping features with SLE [16], [17]that was finally recognized as a distinct clinical entity with specific features such as a relatively good prognosis and response to corticoid treatment [18]. Several sets of diagnostic criteria were suggested with varying sensitivity and specificity [19], [20]. An important biological criterion is the presence of high amounts of anti-ribonucleoprotein antibodies with a particular specificity for components of U1 small nuclear ribonucleoprotein [21].

Our study was conducted on a set of 44 patients that were categorized as SLE (34/44) or MTCD (10/44) on the basis of standard algorithms including clinical and biological criteria. We processed 13 quantitative or categorical biological criteria with currently used machine learning tools in order (i) to assess their capacity to discriminate between SLE and MTCD (supervised classification). (ii) To assess the possibility of reducing the number of parameters without decreasing classification efficiency. (iii) to determine the importance of refining hidden parameters or preprocessing data and (iv) to determine whether SLE and MTCD could appear as separate groups by performing unsupervised classification.

## 2 Methods

### 2.1 Patients and data set

This retrospective study exclusively used data issuing from healthcare and all serum samples were part of a declared Biobank (DC 2012_1704). It was approved and registered by the institution (under GDPR number 20-390) and fulfilled local requirements in terms of data collection and protection of data. Forty four patients were included. Thirty four of them were diagnosed as SLE on the basis of standard criteria [3]. Ten were diagnosed as MCTD on the basis of Alarcon-Segovia criteria [19].

Attempts at classification were performed with 13 parameters that were chosen on the basis of availability and possible relevance to studied diseases. In this first exploratory report, it was not found warranted to attempt at presenting a rationale of this choice and discussing possible confusion between some of the parameters shown below and initial patient classification.

- Age at diagnosis (1) and sex (2).
- Anti-nuclear antibody fluorescence titre (3) and pattern (4) (Kallestad™, Bio-Rad Laboratories, Hercules, CA, USA) as widely used markers of autoimmune rheumatic diseases [22], [23].
- Presence of antibody directed at one or several extractable nuclear antigens among SSA (60kDa), TRIM-21, SSB, SmD, Jo-1, Scl70 and Centromere B (5), as first considered as potential parameters to discriminate between MCTD and SLE [16], and presence and amount of antibodies directed at U1 ribonucleoprotein or specific isoforms such as A, C or 40/43 kD, as detected with EliA™ or Western blotting (Phadia AB, Thermo Fisher Scientific, Uppsala, Sweden) (7-13). Indeed, anti-U1-RNP antibodies are currently used as MCTD markers [19]. A long-term incentive for this choice was a search for a more sensitive marker of MCTD than currently studied anti-U1RNP. As shown below, only anti-43/43kD antibodies were found to contribute discrimination between SLE and MTCD.
- Amount of anti native DNA antibodies, a standard SLE marker (6)

### 2.2 Statistical analysis

Standard statistical analysis was performed with Libre Office statistical tools (*http://www.libreoffice.org*). Comparison between proportions was performed with z test ([24], p125). Comparison between mean values (rough or encoded values) was performed with student’s t-test without using the assumption of equal variances.

Supervised classification and clustering were mostly performed with tools provided by scikit-learn, an open-source machine learning package *(http://scikit-learn.org*). This includes extensive online information. In addition, its use is facilitated by excellent printed tutorials ([25] [26] [27]and it was used in many important studies ([28] [29] are only two examples among many).

The capacity of individual parameters to discriminate between SLE and MCTD was performed with a custom-made python program allowing to compare the composition of the patients groups with respectively lower and higher values than all possible threshold values ranging between the lowest and highest value.

The performance of separation models was systematically assessed with the following methods;

- The *prediction accuracy pa*, i.e. the proportion of exact determinations is widely used but may be deceptive. Thus, if all patients were classified as “SLE”, *pa* would be 38/43 = 0.88, which might seem quite reasonable !
- The *corrected prediction accuracy cpa* (*https://en.wikipedia.org/wiki/Rand_index#Adjusted_Rand_index*) may be viewed as an accuracy score corrected for the occurrence of random guesses according to the formula :

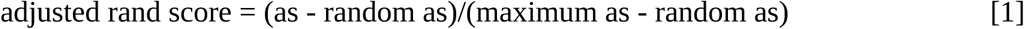

The calculation of this index is implemented in scikit-learn as adjusted rand score. As expected, the calculated would be zero if all patients were classified as SLE.

- A general problem in the elaboration of a binary classifier expected to discriminate between so-called “positive” and “negative” elements is that there is usually a trade-off between the sensitivity, i.e the fraction of positive elements that are classified as positive, and the specificity, i.e. the fraction of negative elements that are classified of negative. The plot of sensitivity versus false positive rate (1 - specificity) is called the ROC curve, and the *area under curve* (*auc*) is often considered as a convenient reporter of the performance of a classifier. An area of 1 corresponds to a perfect classifier, and 0.5 represents a random classifier ([[30], [2]

It well known that the tentative classification of an insufficient number of items based on too numerous parameters may result in deceptive success of a model, a situation denominated as overfitting. The standard way of dealing with this problem consists of splitting a data set beween a training subset, used to calculate classification parameters, and a test subset, used to assess the model performance. Since this procedure may not be sufficient to exclude deceptive conclusions, it may be useful to randomly repeat the splitting process in order to test the variability of performance parameters. This was easily performed in the present study with the train_test_split function of scikit-learn metrics module.

## 3 Results

### 3.1 Potential of single parameters to discriminate between SLE and MCTD

First, we asked which parameters were expected to allow efficient discrimination between SLE and MCTD populations. Five parameters out of 13 yielded different mean values (P<0.05) for both patients’ populations (*Table 1*). For each parameter, we assessed the possibility of separating both populations by using a threshold to define two classes corresponding to lower and higher values. As shown on *Table 1*, the area under roc curve (*auc*) obtained by varying the threshold ranged between 0.596 and 0.835. These values were higher than 0.5, suggesting actual discriminatory power. The maximum prediction accuracy (*pa*) of separation criteria ranged with 0.77 and 0.84. Since these fairly high values were difficult to interpret, a score corrected by tentatively subtracting chance classification success was also used to correct for the possibility of random success in classification. As shown in Table 1, corrected prediction accuracy (*cpa*) ranged between 0.19 and 0.4 for four parameters, thus supporting the hypothesis that the use of a threshold value might provide some discriminatory power.

**Table 1.**
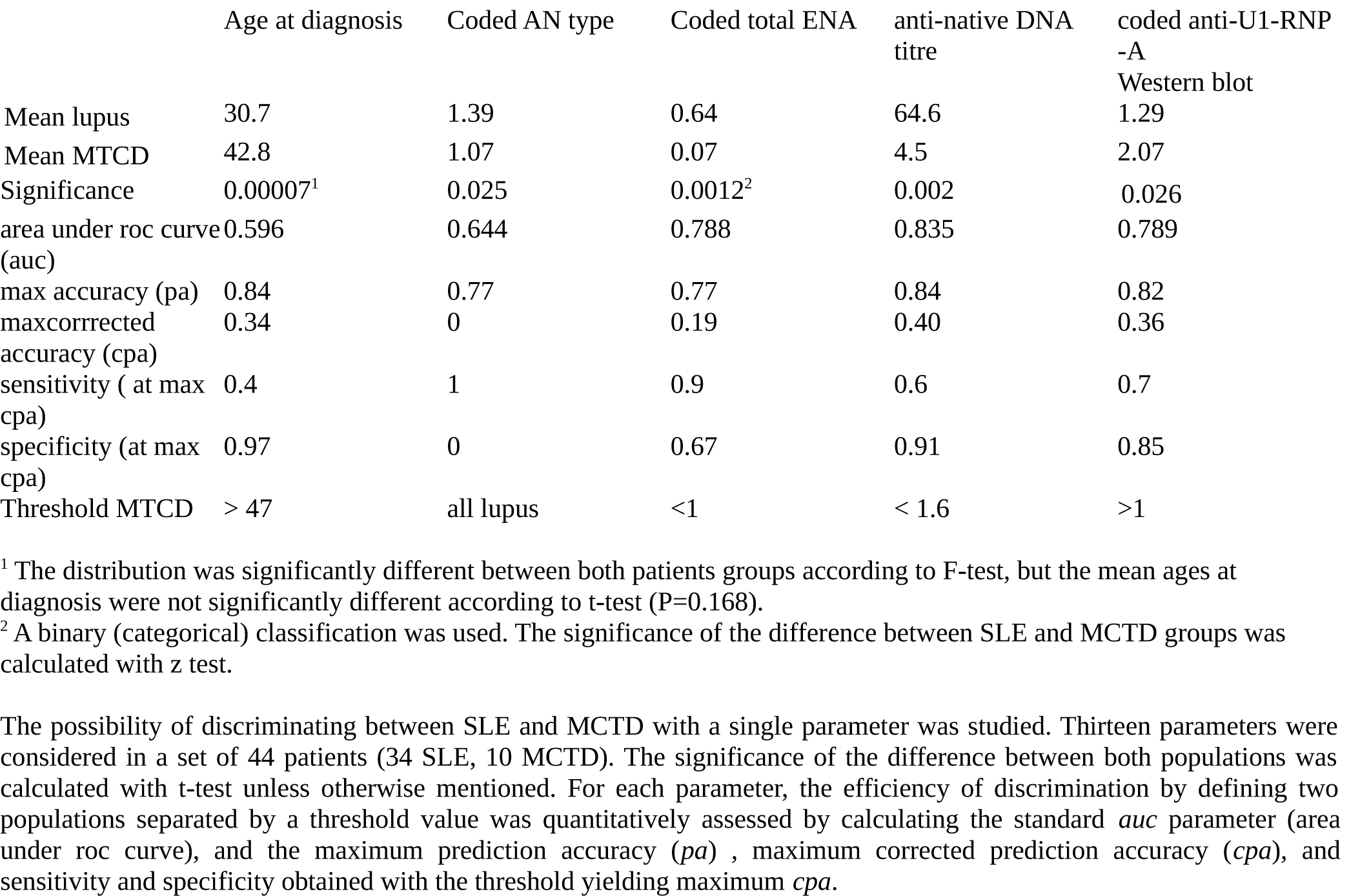
Parameters with significant differences between both patients’ groups.

|  | Age at diagnosis | Coded AN type | Coded total ENA | anti-native DNA titre | coded anti-U1-RNP -A Western blot |
| --- | --- | --- | --- | --- | --- |
| Mean lupus | 30.7 | 1.39 | 0.64 | 64.6 | 1.29 |
| Mean MCTD | 42.8 | 1.07 | 0.07 | 4.5 | 2.07 |
| Significance | 0.00007<sup>1</sup> | 0.025 | 0.0012<sup>2</sup> | 0.002 | 0.026 |
| area under roc curve (auc) | 0.596 | 0.644 | 0.788 | 0.835 | 0.789 |
| max accuracy (pa) | 0.84 | 0.77 | 0.77 | 0.84 | 0.82 |
| maxcorrected accuracy (cpa) | 0.34 | 0 | 0.19 | 0.40 | 0.36 |
| sensitivity (at max cpa) | 0.4 | 1 | 0.9 | 0.6 | 0.7 |
| specificity (at max cpa) | 0.97 | 0 | 0.67 | 0.91 | 0.85 |
| Threshold MTCD | > 47 | all lupus | <1 | < 1.6 | >1 |

<sup>1</sup> The distribution was significantly different between both patients groups according to F-test, but the mean ages at diagnosis were not significantly different according to t-test ( $P=0.168$ ).
<sup>2</sup> A binary (categorical) classification was used. The significance of the difference between SLE and MCTD groups was calculated with z test.
The possibility of discriminating between SLE and MCTD with a single parameter was studied. Thirteen parameters were considered in a set of 44 patients (34 SLE, 10 MCTD). The significance of the difference between both populations was calculated with t-test unless otherwise mentioned. For each parameter, the efficiency of discrimination by defining two populations separated by a threshold value was quantitatively assessed by calculating the standard *auc* parameter (area under roc curve), and the maximum prediction accuracy (*pa*), maximum corrected prediction accuracy (*cpa*), and sensitivity and specificity obtained with the threshold yielding maximum *cpa*.

Interestingly, when the coded fluorescence aspect was studied, *cpa* was often zero, and sometimes exhibited negative values (not shown). This was surprising since the area under roc score (0.644) was markedly higher than 0.5. Also, the distribution of this parameter exhibited highly significant difference between SLE and MTCD, although the average values of this parameter did not significantly differ according to student’s test (P=0.17). A possible interpretation of this apparent discrepancy would be that the distribution of fluorescence aspects might be multimodal in SLE or MTCD.

As an example, the roc curve and threshold dependence of scores is shown on *Figure 1*.

**Figure 1:**
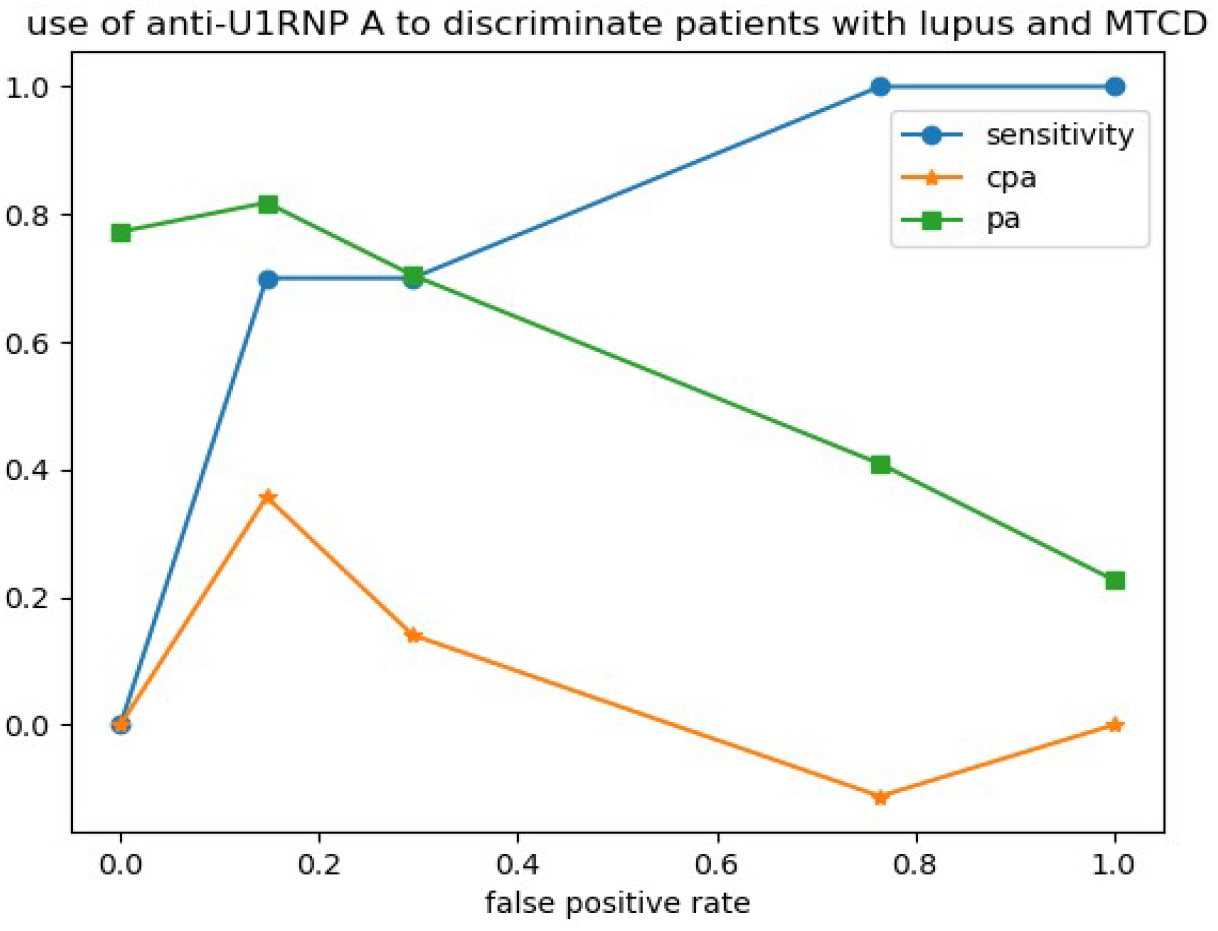
Discrimination between SLE and MCTD on the basis of anti-U1 RNPA

A general problem was that the power of a parameter threshold to separate both populations might be an artifact related to different causes unrelated to the pathological state. A general way to assess this problem consists of splitting the data sets between a training set used to determine the threshold and a test set allowing to assess the validity of this threshold. This possibility was studied for each parameter by performing 25 random splittings to yield a train and a test data set. The average values and standard deviations of area under roc score (*auc*), accuracy score (*pa*) and corrected accuracy score (*cpa*) were calculated. Results are shown on *Table 2*. Interestingly, data supported our previous conclusion and the accuracy scores determined on test sets on the basis of the thresholds calculated on training sets were only moderately decreased as compared to the scores measured on train tests.

**Table 2.** Reproducibility of single parameter discrimination between patients’groups.

| Dataset | Parameter | Age at diagnosis | Coded AN type | Coded total ENA | anti-native DNA title | coded anti-U1-RNP-A Western blot |
| --- | --- | --- | --- | --- | --- | --- |
| train | auc | 0.597 (0.082 SD) | 0.642 (0.037 SD) | 0.787 (0.037 SD) | 0.840 (0.040 SD) | 0.791 (0.047 SD) |
| train | max cpa | 0.360 (0.094 SD) | 0.000 (0.002 SD) | 0.189 (0.070 SD) | 0.416 (0.125 SD) | 0.358 (0.092 SD) |
| train | pa | 0.821 (0.135 SD) | 0.773 (0.042 SD) | 0.726 (0.038 SD) | 0.845 (0.040 SD) | 0.820 (0.034 SD) |
| train | threshold | $\geq 48.1$ (8.8 SD) | $< 0.017$ (0.129 SD) | $< 1$ (0 SD) | $< 1.656$ (1.300 SD) | $\geq 2$ (0 SD) |
| test | cpa | 0.181 (0.284 SD) | 0.025 (0.153 SD) | 0.193 (0.227 SD) | 0.272 (0.312 SD) | 0.320 (0.291 SD) |
| test | pa | 0.780 (0.131 SD) | 0.754 (0.132 SD) | 0.732 (0.115 SD) | 0.805 (0.117 SD) | 0.813 (0.101 SD) |

Patients' data were randomly split between a training data set and a test dataset. The training dataset was used to determine the optimal threshold value allowing to discriminate between SLE and MCTD. The corrected prediction accuracy (*cpa*) and prediction accuracy (*pa*) were calculated for both train and test data sets. The area under roc score (*auc*) was calculated for train dataset. Mean values of 1000 different splits are shown together with standard deviation. Note that standard error was about $(1/1000)^{1/2}$ times the standard deviation.

The general conclusion yielded by these calculations is that all five considered parameters contained significant pieces of information with a potential to discriminate between SLE and MCTD. Obviously, an attractive prospect would be to combine the information contained by these parameters in order to build a more efficient diagnostic model. This was an incentive to explore the potential of currently used machine learning methods to achieve this goal. First, we tred to use available tools with default values of hidden parameters.

### 3.2 Potential of standard machine learning tools to discriminate between SLE and MCTD

First, eight widely used models were used to build classification models on the basis of 13 parameters. A shown on *Table 3*, when models were used with default parameters, the following conclusions were obtained :

- *i*) a number of models were highly efficient in fitting training data sets, yielding up-to 1.000 prediction accuracy or area under roc curve.
- *ii*) Much lower scores were obtained when trained models were used to analyze test datasets. This decrease was much higher than found with aforementioned single parameter classifications.
- *iii*) A substantial variability was found in calculated efficiency indices. Interestingly, additional tests (not shown) revealed that this stemmed from both the variability in the random choice of a training and a test data sets, and in some cases the variability of models involving random procedures. As a consequence, calculations were repeated 25 times for each studied conditions, and mean values are always shown together with standard deviation.

**Table 3.** Efficiency of data analysis with standard methods and settings.

| Method | Training data |  |  | Test data |  |  |
| --- | --- | --- | --- | --- | --- | --- |
|  | pa | cpa | auc | pa | cpa | auc |
| LogisticRegression<sup>1</sup> | 0.936 (0.035 SD) | 0.727 (0.130 SD) | 0.975 (0.016 SD) | 0.727 (0.103 SD) | 0.030 (0.164 SD) | 0.815 (0.140 SD) |
| Support Vector Machine (linear)<sup>2</sup> | 0.961 (0.039 SD) | 0.838 (0.151 SD) | 0.988 (0.015 SD) | 0.742 (0.120 SD) | 0.116 (0.229 SD) | 0.794 (0.123 SD) |
| Support Vector Machine<sup>2</sup> | 0.777 (0.039 SD) | 0.012 (0.057 SD) | 0.830 (0.077 SD) | 0.767 (0.115 SD) | 0.000 (0.000 SD) | 0.578 (0.249 SD) |
| K Neighbors<sup>3</sup> | 0.879 (0.035 SD) | 0.468 (0.185 SD) | 0.923 (0.032 SD) | 0.767 (0.109 SD) | 0.112 (0.233 SD) | 0.828 (0.160 SD) |
| Decision Tree | 1.000 (0.000 SD) | 1.000 (0.000 SD) | 1.000 (0.000 SD) | 0.738 (0.129 SD) | 0.147 (0.243 SD) | 0.616 (0.187 SD) |
| Random Forest | 1.000 (0.000 SD) | 1.000 (0.000 SD) | 1.000 (0.000SD) | 0.727 (0.138 SD) | 0.082 (0.203 SD) | 0.780 (0.156 SD) |
| Gradient Boosting | 1.000 (0.000 SD) | 1.000 (0.000 SD) | 1.000 (0.000SD) | 0.727 (0.123 SD) | 0.074 (0.180 SD) | 0.664 (0.214 SD) |
| Neural Network<sup>4</sup> | 1.000 (0.000 SD) | 1.000 (0.000 SD) | 1.000 (0.000 SD) | 0.763 (0.103 SD) | 0.158 (0.263 SD) | 0.752 (0.141 SD) |

<sup>1</sup> The maximum number of iterations was raised from 200 (default value) to 1000 to achieve proper convergence. Regularization parameter C was set at 1.
<sup>2</sup> The maximum number of iterations was set at 5 000 000 to achieve proper convergence.
<sup>3</sup> Data were preprocessed by scaling all parameters with respect to median value and percentiles (using RobustScaler tool). number of neighbors was set at 5.
<sup>4</sup> Data were preprocessed as was done for KNeighborsClassifier and the number of hidden layers was set at 10 instead of 100 (the default value). The maximum number of iterations was set at 1 000 000.
Standard machine learning tools were used to perform a supervised classification of a data set including 44 patients (34 SLE, 10 MCTD, 13 measured parameters). For all models, the data set was randomly split twenty five times into a training set (33 patients) and a test set (11 patients). Models were trained with the first set using default parameters provided by scikit-learn, unless otherwise mentioned. The prediction accuracy (*pa*), corrected prediction accuracy (*cpa*) and area under roc curve (*auc*) were calculated for the training and test datasets, and mean values are shown together with standard deviation. Note that the expected standard deviation of the mean is (1/5) x SD.

It seemed a reasonable interpretation of these findings to conclude that machine learning models were sufficiently versatile to fit complex data sets, but that the application of these models to test data sets resulted revealed a drastic decrease of classification efficiency, indicative of overfitting. We tested the hypothesis that this overfitting could be reduced by decreasing the number of classification parameters (also called features) and selecting the parameters most relevant to the studied biological problem. This hypothesis was tested by repeating aforementioned calculations to the following conditions : i) a data set restricted to the five most relevant features described in *Table 1*. Results are shown on *Table 4*. ii) a data set restricted to the eight supposed least relevant features. Results are shown on *Table 5*.

**Table 4.** Effect of the reduction of parameter number on classification efficiency.

| Method | Training data |  |  | Test data |  |  |
| --- | --- | --- | --- | --- | --- | --- |
|  | *pa* | *cpa* | *auc* | *pa* | *cpa* | *auc* |
| LogisticRegression<sup>1</sup> | 0.875 (0.038 SD) | 0.505 (0.136 SD) | 0.945 (0.025 SD) | 0.775 (0.093 SD) | 0.137 (0.226 SD) | 0.848 (0.140 SD) |
| Support Vector Machine (linear)<sup>2</sup> | 0.897 (0.033 SD) | 0.589 (0.121 SD) | 0.958 (0.025 SD) | 0.800 (0.025 SD) | 0.243 (0.245 SD) | 0.847 (0.140 SD) |
| Support Vector Machine<sup>2</sup> | 0.775 (0.038 SD) | 0.000 (0.000 SD) | 0.817 (0.067 SD) | 0.767 (0.115 SD) | 0.000 (0.000 SD) | 0.618 (0.196 SD) |
| K Neighbors<sup>3</sup> | 0.885 (0.036 SD) | 0.510 (0.159 SD) | 0.926 (0.034 SD) | 0.775 (0.118 SD) | 0.212 (0.265 SD) | 0.835 (0.158 SD) |
| Decision Tree | 1.000 (0.000 SD) | 1.000 (0.000 SD) | 1.000 (0.000 SD) | 0.731 (0.153 SD) | 0.133 (0.250 SD) | 0.605 (0.191 SD) |
| Random Forest | 1.000 (0.000 SD) | 1.000 (0.000 SD) | 1.000 (0.000 SD) | 0.767 (0.115 SD) | 0.167 (0.230 SD) | 0.863 (0.116 SD) |
| Gradient Boosting | 1.000 (0.000 SD) | 1.000 (0.000 SD) | 1.000 (0.000 SD) | 0.724 (0.127 SD) | 0.087 (0.205 SD) | 0.670 (0.276 SD) |
| Neural Network<sup>4</sup> | 0.992 (0.020 SD) | 0.963 (0.087 SD) | 0.999 (0.003 SD) | 0.764 (0.109 SD) | 0.177 (0.223 SD) | 0.810 (0.154 SD) |

<sup>1</sup> The maximum number of iterations was raised from 200 (default value) to 1000 to achieve proper convergence.
Regularization parameter C was set at 1.
<sup>2</sup> The maximum number of iterations was to 5000000 to achieve proper convergence.
<sup>3</sup> Data were preprocessed by scaling all parameters with respect to median value and percentiles (using RobustScaler tool). number of neighbors was set at 5.
<sup>4</sup> Data were preprocessed as was done for KNeighborsClassifier and the number of hidden layers was set at 10 instead of 100 (the default value). Maximum number of iterations was set at 1 000 000.

**Table 5.** Effect of the use of less relevant parameters on classification efficiency.

| Method | Training data |  |  | Test data |  |  |
| --- | --- | --- | --- | --- | --- | --- |
|  | pa | cpa | auc | pa | cpa | auc |
| LogisticRegression<sup>1</sup> | 0.823 (0.043 SD) | 0.226 (0.167 SD) | 0.766 (0.057 SD) | 0.720 (0.126 SD) | - 0.020 (0.09 SD) | 0.465 (0.240 SD) |
| Support Vector Machine (linear)<sup>2</sup> | 0.829 (0.049 SD) | 0.267 (0.177 SD) | 0.779 (0.063 SD) | 0.698 (0.147 SD) | 0.007 (0.136 SD) | 0.452 (0.242 SD) |
| Support Vector Machine<sup>2</sup> | 0.775 (0.038 SD) | 0.00 (0.000 SD) | 0.773 (0.079 SD) | 0.767 (0.115 SD) | 0.000 (0.000 SD) | 0.518 (0.224 SD) |
| K Neighbors<sup>3</sup> | 0.784 (0.043 SD) | 0.068 (0.095 SD) | 0.811 (0.051 SD) | 0.745 (0.115 SD) | -0.021 (0.059 SD) | 0.483 (0.224 SD) |
| Decision Tree | 1.000 (0.000 SD) | 1.000 (0.000 SD) | 1.000 (0.000 SD) | 0.604 (0.162 SD) | 0.035 (0.176 SD) | 0.511 (0.194 SD) |
| Random Forest | 1.000 (0.000 SD) | 1.000 (0.000 SD) | 1.000 (0.000 SD) | 0.647 (0.134 SD) | 0.003 (0.164 SD) | 0.528 (0.261 SD) |
| Gradient Boosting | 1.000 (0.000 SD) | 1.000 (0.000 SD) | 1.000 (0.000 SD) | 0.629 (0.150 SD) | 0.051 (0.168 SD) | 0.557 (0.236 SD) |
| Neural Network<sup>4</sup> | 0.972 (0.033 SD) | 0.881 (0.138 SD) | 0.992 (0.012 SD) | 0.585 (0.124 SD) | -0.023 (0.091 SD) | 0.439 (0.225 SD) |

<sup>1</sup> The maximum number of iterations was raised from 200 (default value) to 1000 to achieve proper convergence. Regularization parameter C was set at 1.
<sup>2</sup> The maximum number of iterations was to 5000000 to achieve proper convergence.
<sup>3</sup> Data were preprocessed by scaling all parameters with respect to median value and percentiles (using RobustScaler tool). number of neighbors was set at 5.
<sup>4</sup> Data were preprocessed as was done for KNeighborsClassifier and the number of hidden layers was set at 10 instead of 100 (the default value). Maximum number of iterations was set at 1 000 000.

In view of the complexity of results displayed on *Tables 3, 4*, and *5*, it was deemed warranted to first examine the relationships between the three parameters used to assess the efficiency of different models. First, the correlation between the mean values of *pa, cpa* and *auc* determined under 48 different conditions (25 calculations each) was studied :

- i) parameter *pa* was not significantly corrrelated to *cpa* (r = 0.032, P>0.1) nor to *auc* (r = 0.100, P>01).
- ii) *cpa* and *auc* were strongly correlated (r= 0.826, P<0.01), as shown on *Figure 2*:

**Figure 2:**
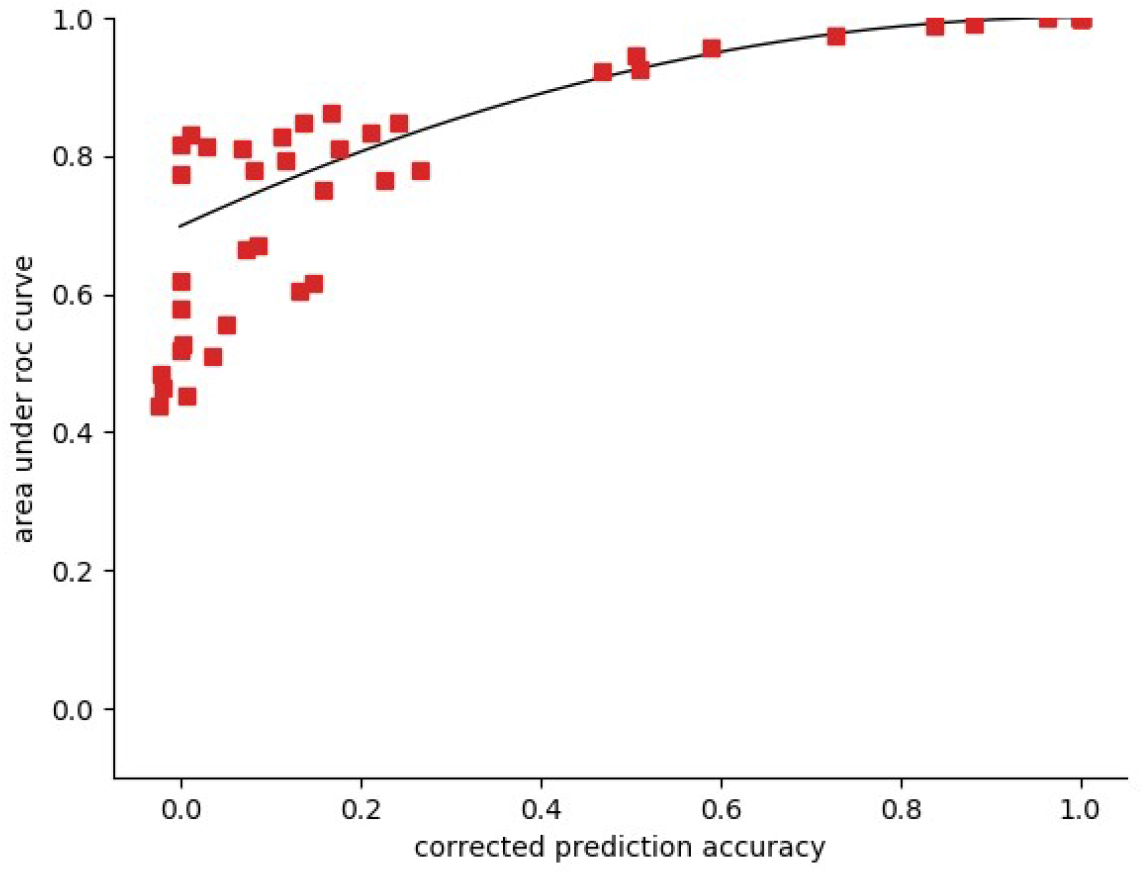
correlation between cpa and auc. Parameters cpa and auc were determined under 48 different conditions (varying machine learning models and data sets) with 25-fold determination. Mean values are shown together with the parabolic regression curve (*auc = - 0.60446 cpa2 + 0.98544 cpa + 0.61182)*.

Since the fairly high values of *pa* may be deceptive when limited asymmetric data sets are studied (indeed, if 90% of patients are classified as 1, a model classifying all patients as 1 would yield a predictive accuracy of 90%), it was deemed reasonable to retain the *cpa/auc* couple as a convenient reporter of classification efficiency.

Parameter *cpa* may convey an intuitive feeling for the quality of a model, with a value of zero in absence of any predictive capacity and 1 in case of perfect match. Parameter *auc* is more widely used in the scientific literature. The interpretation is somewhat more subtle, and a high *auc* value may be indicative of the possibility to adapt the sensitivity/specificity of a given model to medical needs (e.g. by chosing high sensitivity to detect potentially severe and curable conditions).

Thus, we chose to consider only *cpa* and *auc* in the reminder of this study. The numerical results displayed on *Tables 3, 4* and *5* are illustrated on *Figure 3*.

**Figure 3.**
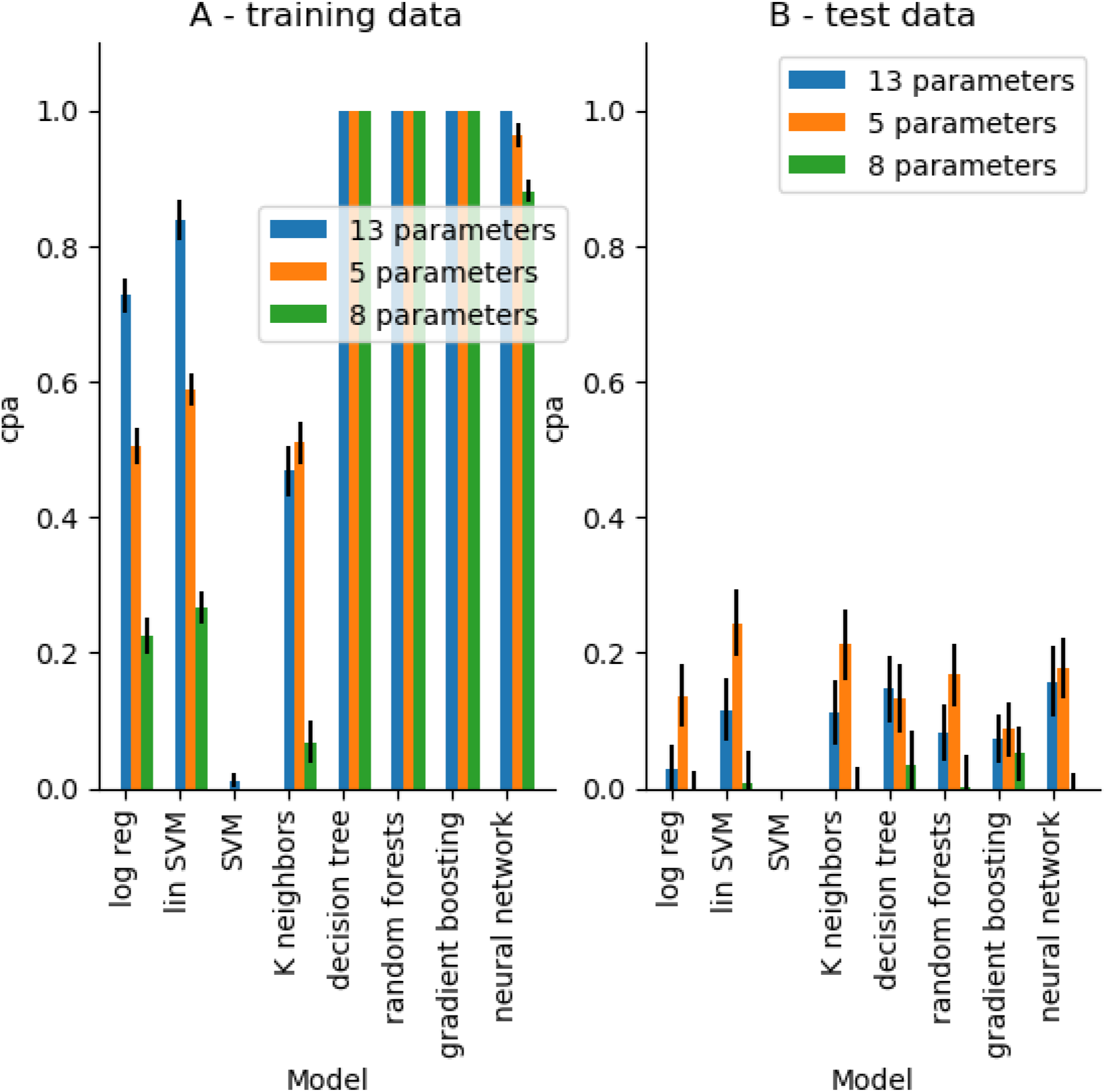
Effect of dataset of model efficiency as assessed with cpa. Eight widely used machine-learning classifiers were used to discriminate between SLE and MTCD on the basis of a dataset including 44 patients and 13 parameters. Caculations were performed with i) all 13 parameters, ii) five parameters,with significant differences between both patients groups, iii) the remaining 8 parameters. The mean cpa obtained after 25 calculations is shown for each model. Vertical bar length is twice the standard error of the mean.

The following conclusions were suggested :

i. Not surprisingly, combining all 13 parameters allowed more efficient fit between models and data than the use of a single parameter described on *Table 2*.
ii. When calculations were based on the 5 supposed most relevant parameters, fitting efficiency was decreased on training data sets, but it was increased on test data sets. This is illustrative of the well known overfitting situation.
iii. when 8 supposed least relevant parameters were used, fitting efficiency was decreased in both training and test data sets. Note that the efficient fitting of training data found with some machine learning models with nearly zero predictive accuracy on test data is a clear illustration of overfitting. This emphasizes the prominent importance of the selection of data set parameters (or features).
iv. The processing of five supposedly relevant parameters did not result in marked improvement of prediction accuracy as compared to single parameter analysis (as shown on *Table 2* and *Table 4*).

The simplest interpretation of these results would be that the low efficiency of machine learning models might be due to a poor relevance of default settings to the properties of the data sets we used. It was thus warranted to study the efficiency increase that might be obtained by a proper setting of the hidden parameters of all classifiers. A number of parameters were thus subjected to systematic variation to address this question. Results are displayed in the following section.

### 3.3 Improving classifier settings

The eight models considered above were sequentially considered for parameter optimization. The five parameter data set defined above was used for all studies.

#### 3.3.1 Logistic Regression

This methods consists of looking for a linear combination of provided parameters allowing optimal discrimination between both patient’s classes. While this is close to standard statistical reasoning, this involves a regularization procedure allowing to achieve a balance between the complexity and efficiency of trained models. A regularization parameter allows to modulate the relative importance of regularization and classification efficiency.

The dependence of *cpa* and *auc* on regularization is shown on *Figure 4*. As expected, the fitting efficiency of training data was increased when regularization was decreased. However, *cpa* and *auc* yielded by the analysis of test data sets were nearly optimal when the regularization parameter was set to 1. Interestingly, this is the default value of the algorithm provided by scikit-learn.

**Figure 4.**
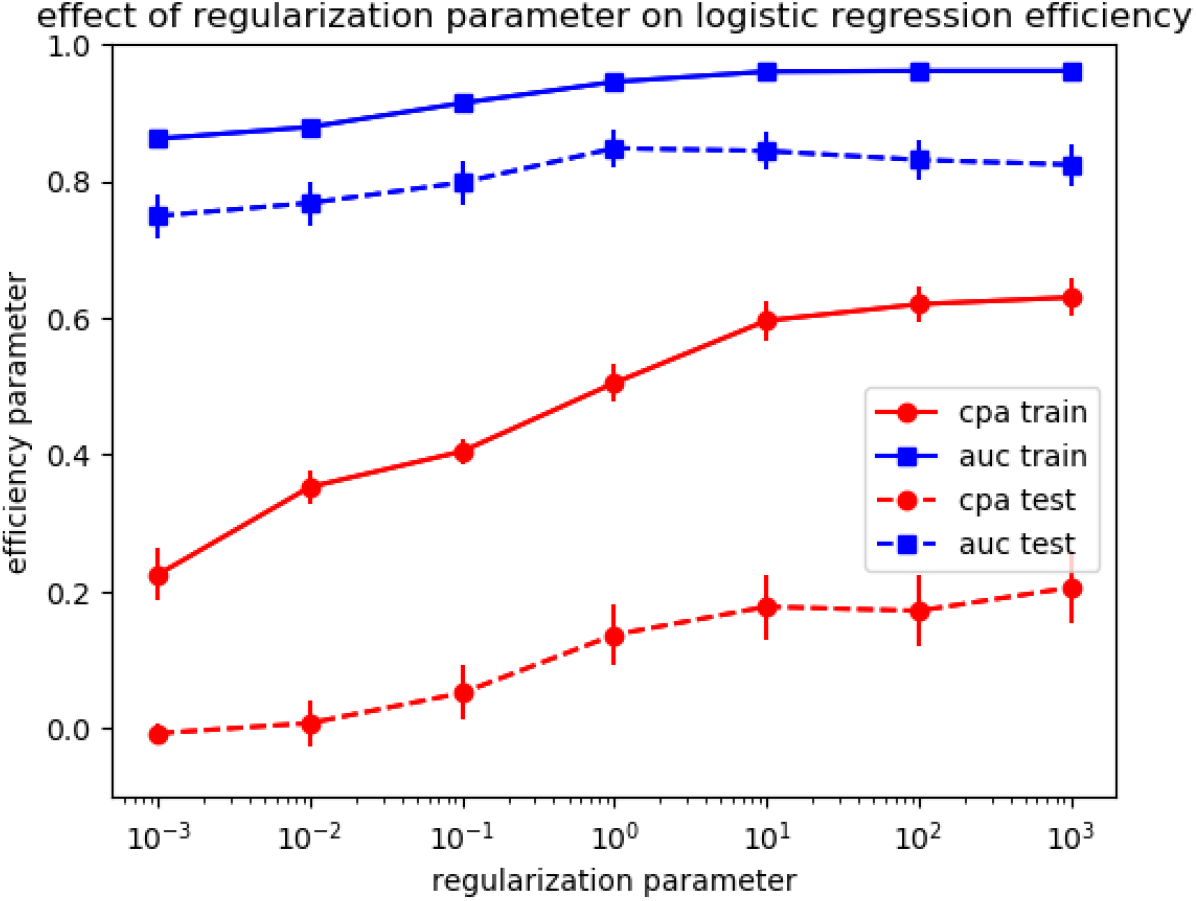
Effect of regularization on logistic regression efficiency. (Note that the relative importance of regularization is inversely related to the regularization parameter). Mean values of 25 sequential determinations are shown for each condition. Vertical bar length is twice the standard error of the mean.

#### 3.3.2 Linear support vector machine classifier

As shown on *Figure 5*, the efficiency of linear SVM and effect of regularization were comparable to those of logistic regression.

**Figure 5.**
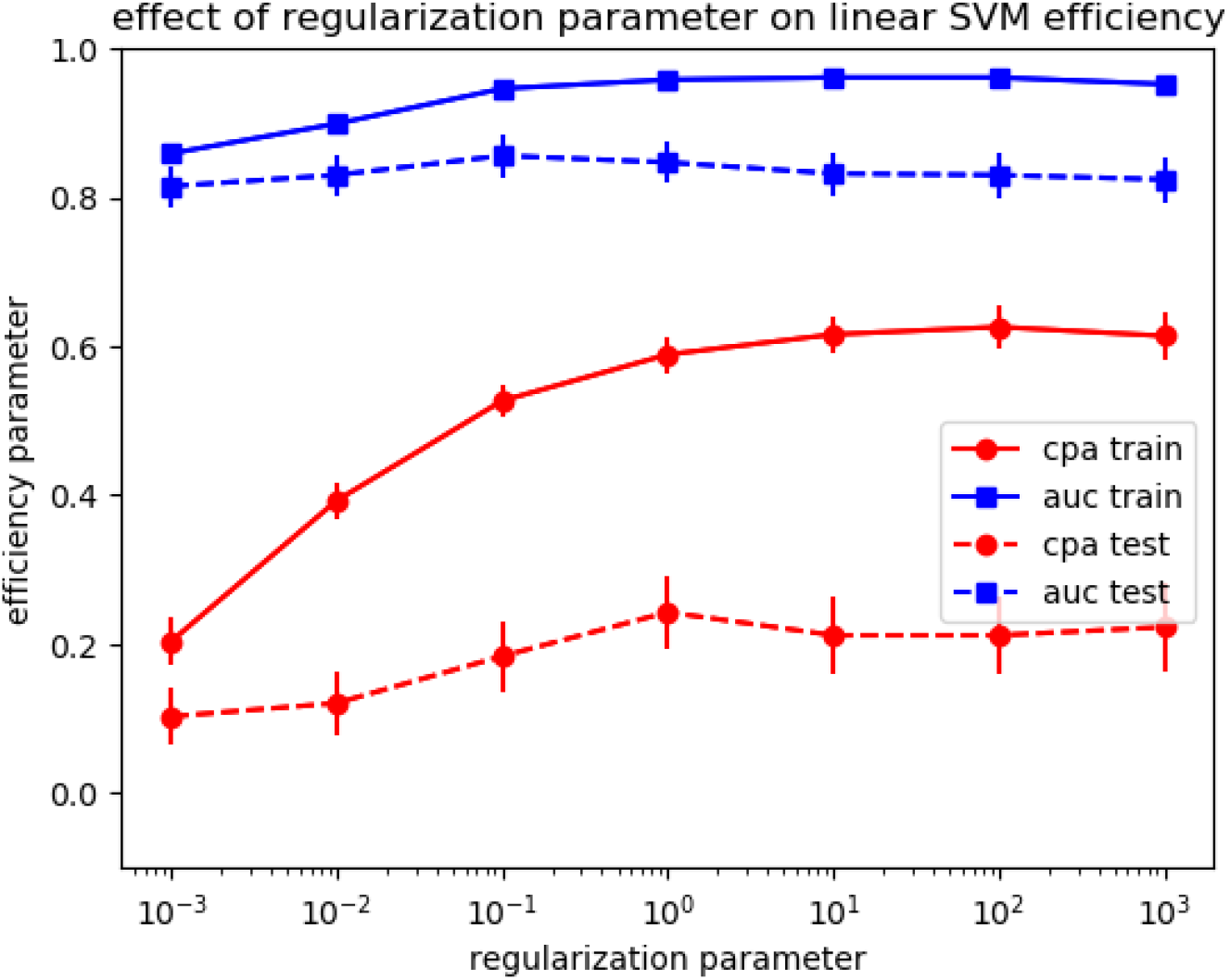
Effect of regularization on linear SVM. (Note that the relative importance of regularization in inversely related to the regularization parameter). Mean values of 25 sequential determinations are shown for each condition. Vertical bar length is twice the standard error of the mean.

#### 3.3.3 Support vector machine

The SVM model may be viewed as an extension of linear SVM with increased versatility. When the value of two hidden parameters, the regularization parameter C, in analogy to the previous two models, and the kernel coefficient *gamma* were varied, the behavior of *cpa* and *auc* as determined for training and test data sets were fairly complex and highest values of each parameter were obtained for different combinations of C and *gamma* : with training data set, high values of *cpa* and *auc* (respectively 0.993 and 1) were obtained with C = 1 (the default value) and *gamma* = 100. However, applying the trained model to test data sets yielded disappointingly low values of *cpa* (0) and *auc* (0.649), suggesting strong overfitting. Further, a fairly different combination (C=10 and *gamma*=0.1) yielded low *cpa* (0.747) with training data set while the analysis of test data sets yielded better values in comparison with previous models (*cpa*=0.264 +/- 0.3 SD and *auc*=0.85 +/- 0.14 SD). In conclusion, results were more critically dependent on the precise values of hidden parameters, thus questioning the robustness of the model.

This conclusion is illustrated on *Figure 6* where the dependence of *auc* (test dataset) on two parameters is shown.

**Figure 6:**
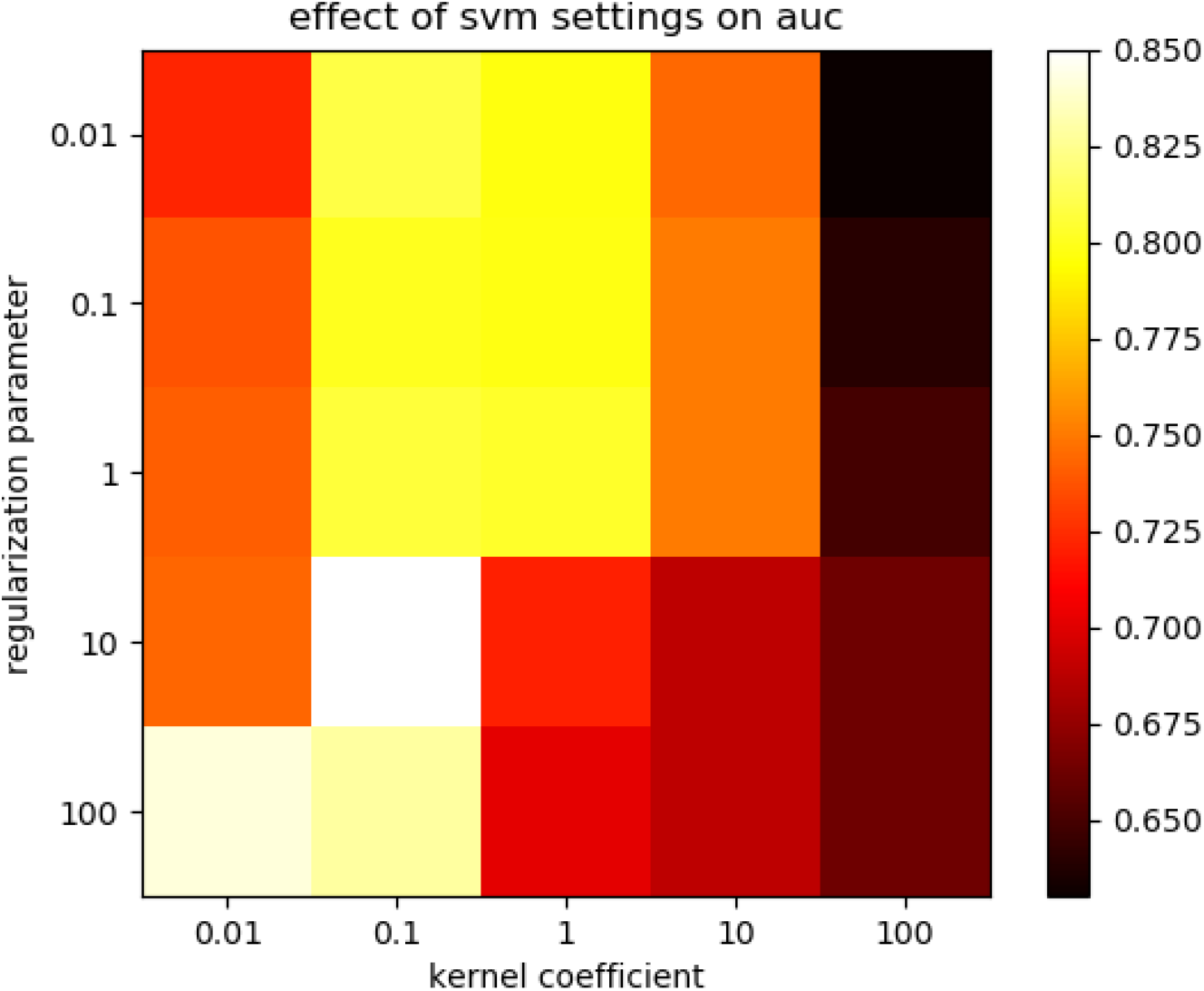
Influence of kernel coefficient and regularization parameter of the classification efficiency of SVM model.

#### 3.3.4 KNeighborsClassifier

The basic principle of this classifier is fairly straightforward: The class of any point (in a space the dimension of which is equal to the number of parameters) is determined as the class of the majority of the *n* nearest points belonging to the training data set. Clearly, if *n* is set to 1, a perfect fit will be obtained for all points of the training data set. If n is set equal to the number of points in the data set, all points will be classified according to the most populated class in the data set. The dependence of classification efficiency on n is shown on *Figure 7*.

**Figure 7.**
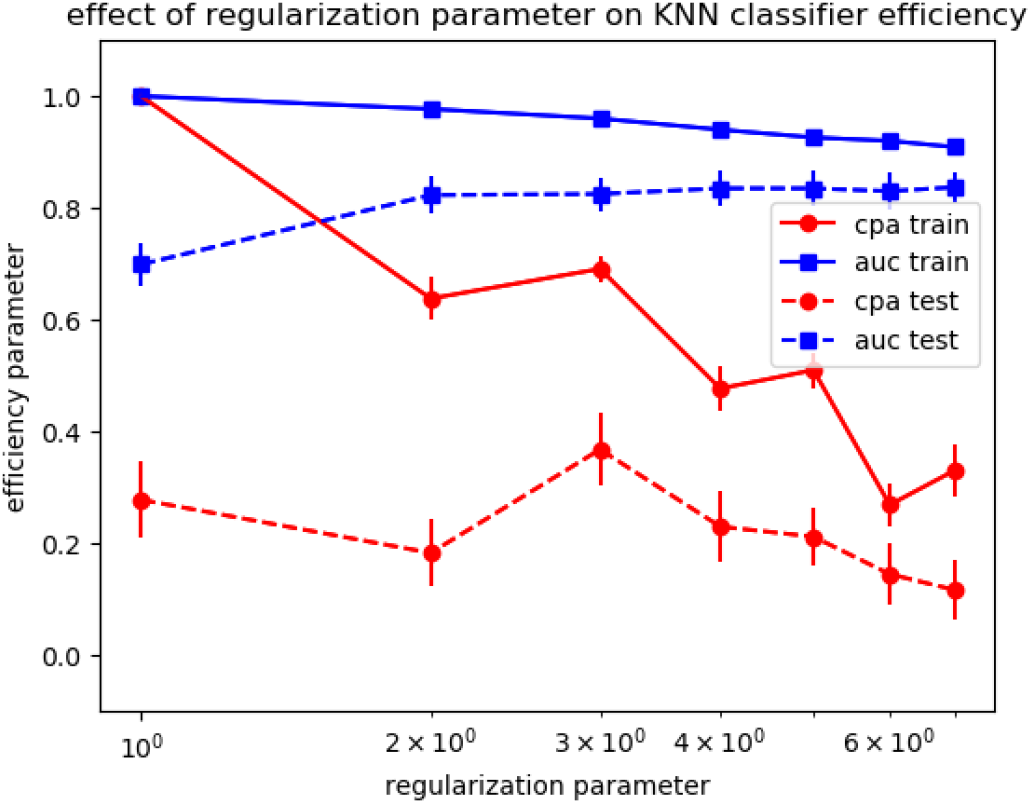
Importance of the choice of neighbor number on the efficiency of KNN classifier. Each point represents the mean (+/- Standard error) of 25 calculations based on the same parameters, with random determination of training and test data sets. Distance was calculated on the basis of parameters scaled with RobustScaler method provided by ScikiLearn.

As shown on *Figure 7*, decreasing the number of neighbors from 5 (the default value) to 3 improved *cpa* and *auc*. An important point is about the definition of the distance between data points : This is obviously strongly dependent on the scaling of parameters. Thus, if a parameter is expressed as concentration displaying 1000 fold variation and another parameter as an encoded value with a range of 1-5, the first parameter is expected to play a dominant influence on calculated distance. Therefore, an appropriate scaling must be performed to balance the relative importance of parameters.

Note that specific scaling procedures inspired by some biological reasoning might be expected to improve classification efficiency. Addressing this point was not felt to fit to the scope of the present report.

#### 3.3.5 Importance of the maximum depth of decision tree classification

The basic principle of this method is fairly simple : this consists of performing sequential binary splittings until an optimal fit is obtained. Clearly, if the maximum number of allowed splits is unlimited, a perfect fit is expected to be obtained with any data set. However, if parameters are irrrelevant to classification, this should result in bad accuracy of test data set analysis. The dependence of classification efficiency on the maximum number of allowed splits (max_depth parameter) is shown on *Figure 8*.

**Figure 8.**
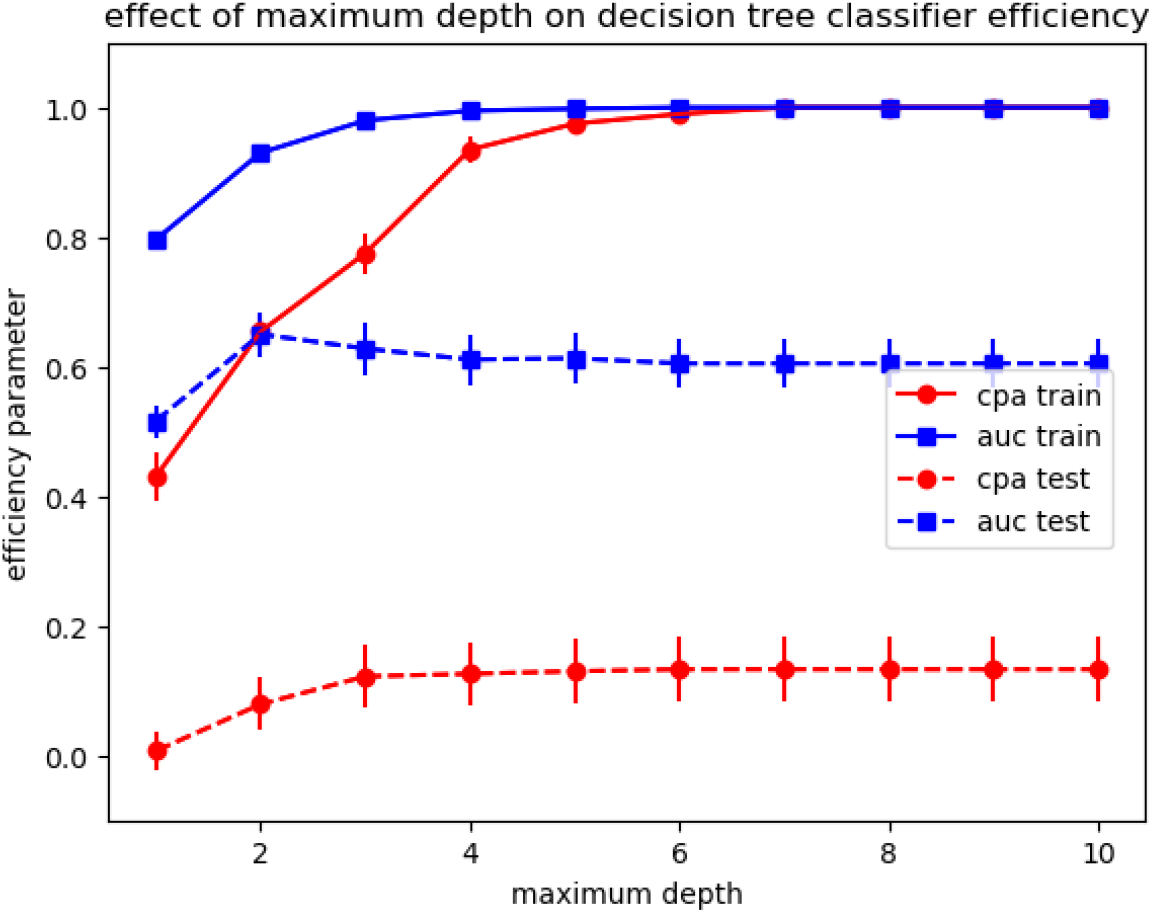
Importance of the choice of maximum depth parameter on the efficiency of decision tree classifier. Each point represents the mean (+/- Standard error) of 25 calculations based on the same parameters, with random determination of training and test data sets.

Clearly, while the accuracy of test data set analysis was somewhat improved by a reduction of the maximum depth, the decision tree algorithm yielded poor accuracy parameters as compared to other methods, despite an efficient fitting with training dataset (*Table4*).

#### 3.3.6 Random forests classifier

The random forest method may be viewed as an extension of the decision tree classifier consisting of combining the data obtained by several decision tree calculations differing with the random choice of sequential data set splittings during the training process. The maximum *cpa* value was 0.185 +/- 0.275 SD, corresponding to a nearly maximum value of 0.868 +/- 0.123 SD for *auc*.

The dependence of *auc* (test dataset) on varied parameters is displayed on *Figure 9*.

**Figure 9.**
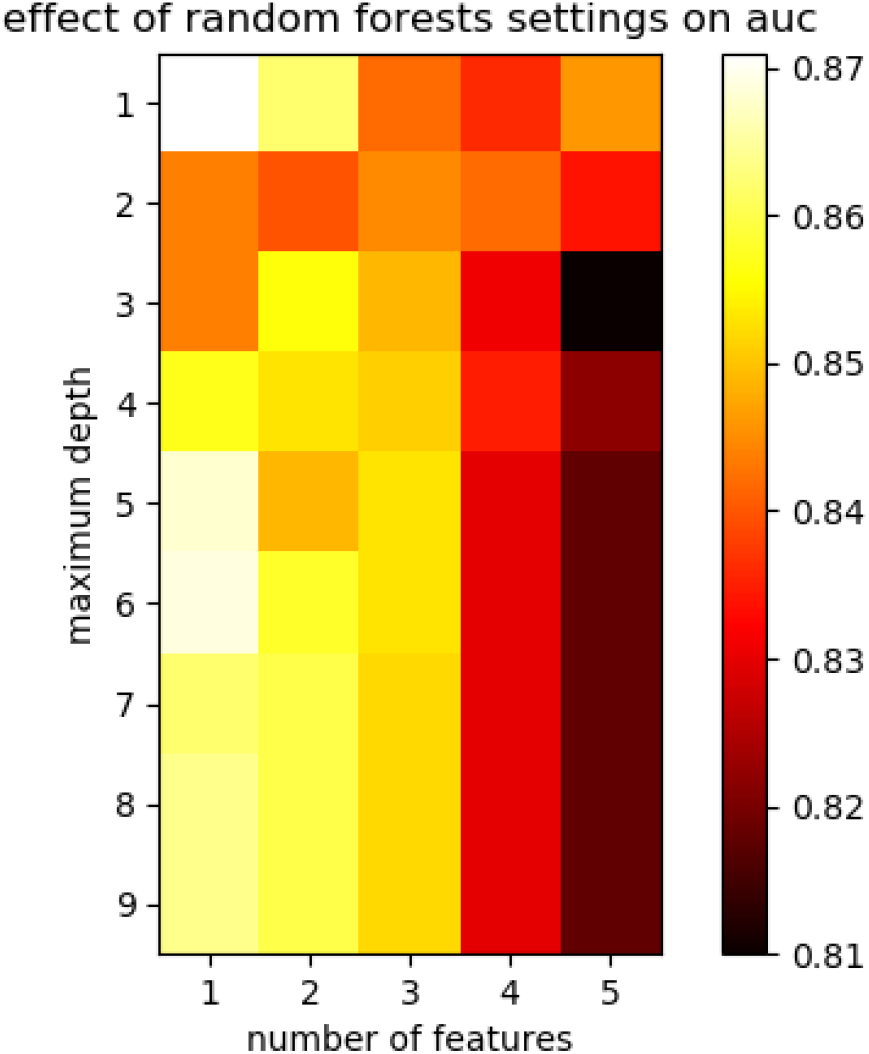
Influence of maximum depth and number of feature on random forest classifier efficiency. The random forest classifier algorithm was used with varying values of maximum_depth and number of features (max_feature) parameters. The number of combined trees was set at 200. Each calculation was repeated 25 fold under each condition and mean values of parameters *cpa* and *auc* obtained on test data set with models trained on training data set were calculated.

#### 3.3.7 Gradient boosting classifier

The gradient boosting classifier may be viewed as a more complex way of building on the decision tree algorithm with a number of adjustable parameters, including so-called learning_rate parameter allowing to weight the importance of combined estimators. The effect of combined variation of learning_rate and maximum depth is shown on *Figure 10*.

**Figure 10.**
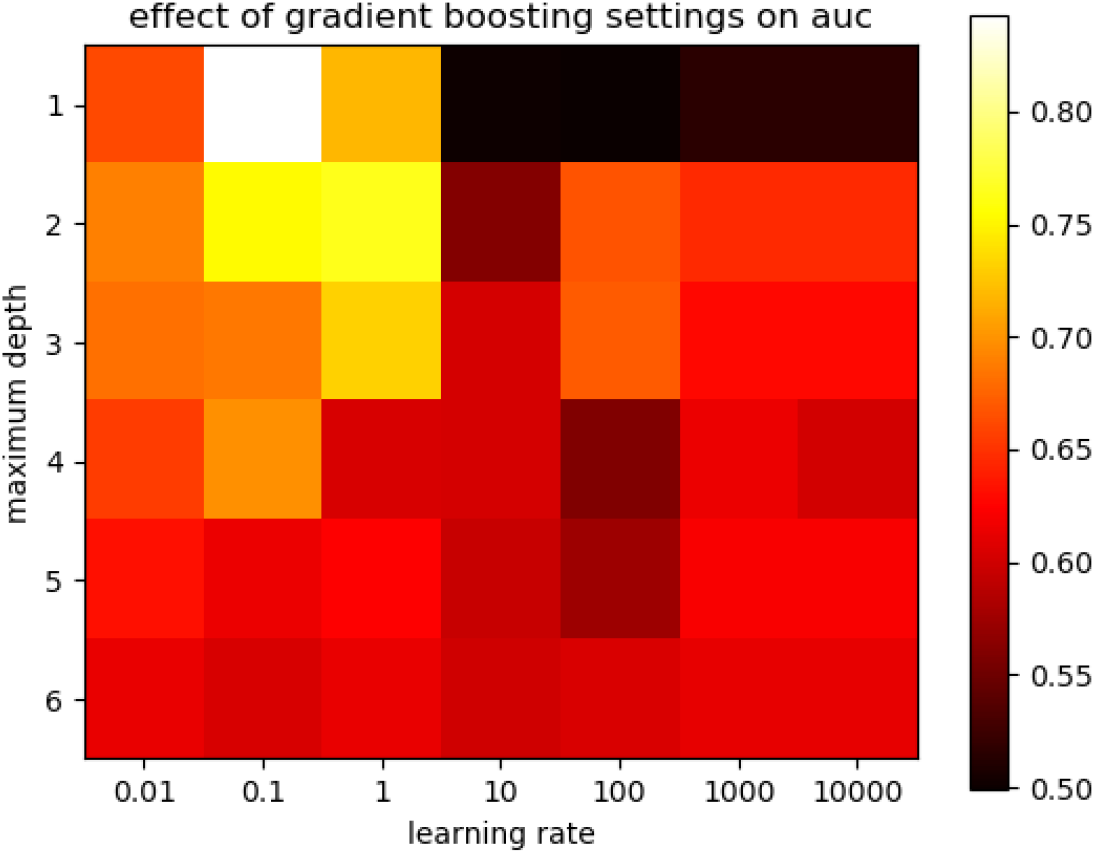
Influence of maximum depth and learning rate on gradient boosting classifier efficiency. The gradient boosting classifier algorithm was used with varying values of maximum_depth and learning rate parameters. The number of combined trees was set at 50. Each calculation was repeated 25 fold under each condition and mean values of parameters cpa and auc obtained on test dataset with models trained on training dataset were calculated.

The maximum value of *cpa* (applied to test data sets) was 0.189 +/- 0.269 when maximum_depth was set at 2 and learning rate at 100, while *auc* was 0.668 +/- 0.209 SD, a fairly low value.

The maximum value of *auc* was 0.843 +/- 0.131 SD when maximum depth was 1 and learning rate 0.1. Under these conditions, parameter *cpa* was 0.148 +/- 0.271 SD.

#### 3.3.8. Neural Networks

Neural networks are highly complex algorithms that gained some popularity several decades after initial reports due to the increasing availability of large scale data sets ([31], [32]. The principle consists of subjecting input parameters to sequential mathematical transformations within so-called hidden layers, resulting in enormous complexity and versatility. Due to the high number of adjustable parameters, this model is probably poorly suited to the analysis of small data sets. However, it was felt warranted to present a limited view of its potential.

Optimal fit yielded reasonably high values of 0.316 +/- 0.322 SD for *cpa* and 0.842 +/- 0.126 SD for *auc*.

#### 3.3.9. Section summary

The results presented in this section support the following conclusions.

i. As suggested by *Figure 3*, the selection of parameters should be subjected to careful analysis and limitation before applying powerful machine learning tools.
ii. When only a limited data set is available, the use of more complex algorithms is not advisable, as exemplified on *Table 4*.
iii. A common problem in the analysis of small data sets is the high variability of calculated parameters, due to the occurrence of a number of random steps in currently available algorithms.
iv. All models include a number of adjustable parameters the determination of which is somewhat hampered by random variability.

These provisional conclusion suggested to refine parameter selection and apply less complex models, such as Logistic Regression, linear Support Vector machine, Nearest neighbor analysis or decision trees with limited depth to the analysis of smaller sets of parameters.

### 3.4 Tentatively reducing the number of parameters

In order to determine the optimal parameter combination for patient classification, we tested all subsets of 2 parameters (10 possible choices), 3 parameters (20 possibilities) or 4 parameters (5 possibilities) that could be selected out of the five most significant parameters shown on *Table 1*. In each case, we studied the efficiency of three machine learning models to classify patients. One thousand combinations of training and test subsets were used and the mean value of corrected prediction accuracy (*cpa*) was determined. The parameter subsets yielding maximum *cpa* are shown on *Table 6*, leading to the following conclusion:

**Table 6.** effect of parameter selection on classification efficiency

| Parameters used | corrected prediction efficiency |  |  |
| --- | --- | --- | --- |
|  | Logistic regression | K nearest neighbors<sup>°</sup> | Decision tree |
| B D | 0.369* +/- 0.011 SE | 0.269 +/- 0.009 SE | 0.300* +/- 0.018 SE |
| A D | 0.189 +/- 0.017 SE | 0.334* +/- 0.010 SE | 0.271 +/- 0.017 SE |
| B C E | 0.389* +/- 0.011 SE | 0.293 +/- 0.010 SE | 0.330* +/- 0.010 SE |
| A B E | 0.130 +/- 0.008 SE | 0.409* +/- 0.011 SE | 0.183 +/- 0.080 SE |
| B C D E | 0.262* +/- 0.009 SE | 0.292 +/- 0.010 SE | 0.023 +/- 0.009 SE |
| A B D E | 0.199 +/- 0.008 SE | 0.358* +/- 0.010 SE | 0.142 +/- 0.016 SE |
| A B C E | 0.242 +/- 0.010 SE | 0.339 +/- 0.010 SE | 0.295* +/- 0.019 SE |

<sup>°</sup> The number of neighbors was set at 3 and parameters were scaled as mentioned on Table 3.
\* This is the maximum efficiency value obtained by all permutation of five parameters shown in Table 1.

i. for a given parameter subset, different models may exhibit up to threefold differences in classification efficiency (thus maximum *cpa* obtained with a three-parameter subset including age at diagnosis, coded anti-nuclear antibody aspect and coded anti-U1-RNP-A assayed with western blot, varied between 0.130 (logistic regression) and 0.409 (K nearest neighbors classifier)
ii. when all results obtained with different methods and parameters were considered (as shown on *Tables 1 & 6*), the maximum *cpa* obtained on test data was 0.320 (+/- 0.010 SE) with 1 parameter, 0.334 +/- 0.010 with 2 parameters, 0.409 +/- 0.011 with 3 parameters and 0.358 +/- 0.01 with 4 parameters. This supported the conclusion that better results were obtained with limiter parameter subsets. Also, the gain provided by the combination of several parameters remained fairly modest.

### 3.5 Prediction probability

Since classifiers usually involve an estimator of the probability of each prediction, it was interesting to examine the estimated probability of aforementioned predictions. Indeed, it might be hypothesized that false predictions might be due to less representative patients, leading to lower prediction probability. Results obtained with three parameter sets are displayed on *Table 7*. While the estimated probability of false predictions was significantly lower than that of true predictions, the probability parameter exhibited too high variations to be used as a warning signal.

**Table 7.** Estimated probability of classifier prediction

| parameters | Estimated prediction probability |  |  |  |  |  |
| --- | --- | --- | --- | --- | --- | --- |
|  | Logistic Regression |  | K Neighbors |  | Decision Tree |  |
|  | true predictions | false predictions | true predictions | false predictions | true predictions | false predictions |
| A,B,E | 0.827 (0.143 SD) | 0.722 (0.148 SD) | 0.900 (0.153 SD) | 0.818 (0.170 SD) | 0.913 (0.153 SD) | 0.834 (0.140 SD) |
| B, C, E | 0.805 (0.115 SD) | 0.669 (0.099 SD) | 0.882 (0.159 SD) | 0.806 (0.164 SD) | 0.819 (0.189 SD) | 0.766 (0.174 SD) |
| A, B, C, D, E | 0.888 (0.155 SD) | 0.747 (0.137 SD) | 0.892 (0.156 SD) | 0.740 (0.138 SD) | 0.973 (0.085 SD) | 0.943 (0.109 SD) |

### 3.6 Tentative unsupervised clustering

A potentially useful point would be to determine whether the studied group of 44 patients displayed some unexpected heterogeneity that might hamper the discrimination between SLE and MCTD. We addressed this point by performing unsupervised clustering to define two groups with KMeans, a commonly used clustering algorithm based on euclidean distance between properly scaled data points. As expected due to the low value of *cpa*, the processing of 5- or 13-parameter data sets yielded a classification that was unrelated to the SLE/MCTD classification, with a corrected matching index close to zero.

## 4 Discussion

Our study was aimed at determining whether currently available machine learning tools might help us processing biological data used to manage patients with connective tissue diseases. Presented results might be useful in two domains:

i. providing caveats and guidelines for the use of machine learning tools. Indeed, as recently emphasized, artificial intelligence is becoming easier and easier to use, with a substantial risk of reaching erroneous conclusions[33].
ii. improving the management of patients with autoimmune diseases.

### 4.1 Potential and limitation of the use of standard machine learning tools to process limited data sets

While the guidelines and caveats illustrated by our results are well known from data scientists, it was felt that the detailed numerical data provided in this study might be of interest for more medically inclined readers. The following five points may be emphasized :

#### 4.1.1 Importance of random variations

Results yielded by calculations are subject to random variations due to both random steps of machine learning algorithms and dependence of data on random properties of data sets, that are the more important as their size is smaller. This is well illustrated by standard deviations displayed on *Table 3*. This is the reason why repeated processing of training and test data sets is an absolute requirement to build models and validate them with independent data sets.

#### 4.1.2 Danger of using too many parameters (or features)

While artificial intelligence met with spectacular success by processing highly complex data sets the relevance of which is sometimes difficult to assess, it must be understood that the price to be paid for these successes is that they may need more data than conventional methods, a property that was sometimes dubbed “data hungriness”. This point is well illustrated on *Figure 3*. It must be emphasized that *i)* this danger is somewhat alleviated by so-called regularization techniques included in machine learning algorithme, and *ii)* a useful warning is provided by the occurrence of an important difference between the classification efficiency of training and test data sets. Data shown on *Table 6* are indicative of the interest of using a very limited number of parameters when small-size data sets are used.

#### 4.1.3 Importance of using simpler models when data sets are smaller

As illustrated on *Figures 4 to 11*, a reason for the simplicity of use of many currently available machine learning tools is that they include cleverly chosen default values of a number of fairly hidden parameters. A simple example is the maximum depth of decision tree classifier, i.e. the maximum number of patient splitting into two categories. Clearly, the use of a number n of splittings result in the definition of 2^n^ classes. As an example, performing 10 splittings will generate about 1,000 classes, which will allow perfect, although fairly meaningless, classification of a few tens of patients. This illustrates the importance of decreasing model complexity by decreasing parameters such as maximum depth and increasing the importance of regularization.

**Figure 11.**
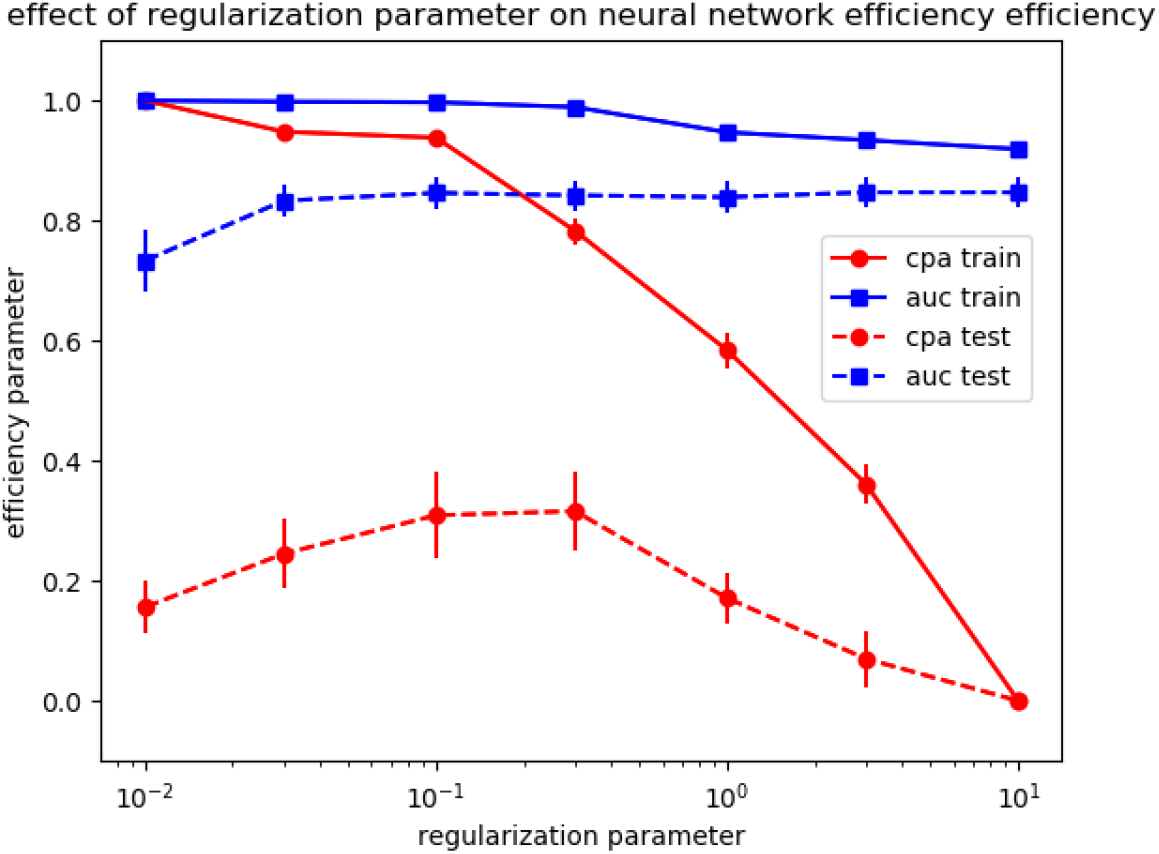
Influence of regularization parameter on multilayer perceptron classification efficiency. The multilayer perceptron classifier algorithm was used with varying values of retularization parameter The number of hidden layers was set at 10. Data transformation was performed with hyperbolic tangent (tanh). maximum iteration was set at 50 000.

#### 4.1.4 Importance of a quantitative assessment of classification efficiency

The impressive number of options concerning the choice of models, model settings and classification features made it clear that there is a need for guidelines to chose appropriate strategies. Model evaluation is indeed a common challenge, and two main parameters were considered in the present report. The corrected prediction accuracy if fairly intuitive, with a value usually ranging between zero, for a totally inefficient algorithm and 1, corresponding to perfect classification, and possibly negative values for models performing still less efficiently that random choices. The meaning of the area under Roc curve is somewhat more subtle, and reminds us of the balance between the sensitivity and the specificity of a test. Also, it cannot be used to assess the efficiency of classification between more than two states. The close correlation between *auc* and *cpa* displayed on *Figure 2* suggests that we may tentatively chose the parameter that suited us best.

#### 4.1.5 Prominent importance of parameter selection

While impressive and unexpected correlations between different features were sometimes obtained by processing massive data sets, results displayed on *Figure 3* clearly support the conclusion that parameter selection is essential when limited data sets are available, as exemplified in the present study. Indeed, different models described above displayed fairly comparable efficiency (*Table 6*). Thus, calculating parameters such as *cpa* and *auc* may help us assess the quality of model selection and setting. As will be emphasized below, machine learning may thus not only facilitate the use of complex diagnostic algorithms but also provide a useful help in defining pathological categories.

### 4.2 Potential use of our study to the discrimination between SLE and MCTD patients

As emphasized above, the continual increase of the diversity, and cost, of diagnostic and therapeutic tools results in a parallel increase of more and more accurate diagnosis. Since pathognomonic signs are rarely available for this purpose, the complexity of diagnostic algorithms is steadily increasing. The discrimination between SLE and MCTD provides a clearcut illustration of this situation.

SLE is a severe chronic disease requiring therapeutic choices for optimal efficiency and minimal adverse effects. MCTD was suggested more than 30 years ago to represent a separate entity that was notable for a lack of renal disease and excellent response to corticosteroid therapy [16]. Twenty years later, the existence of MCTD as a separate entity remained subject to discussion [[18], and diagnostic algorithms including more than 10 biological and clinical criteria were suggested [19]. Clearly, while no definitive procedure is currently available to answer without any ambiguity all questions concerning individual patients, data obtained in the present study suggest that currently available machine learning tools might help us address the following points:

i. estimate the probability that a patient’s pathology might be classified as SLE or MCTD. As shown on *Table 4, auc* parameters as high as 0.84-0.86 might be obtained with a 5-parameter set. An *auc* of 0.827 could be obtained with a 3-parameter set as discussed on *Table 6* (not shown). These values may be considered as fairly good according to current standards [[34]. Indeed, as a recent example, SVM was used to predict the cognitive decline of Parkinson patients subjected to brain fluorodeoxyclucose PET scans. An *auc* of 0.73 was obtained with a model trained on 43 patients [35]. It must be emphasized that more efficient classifications may be obtained on simpler models. Thus, an image analysis software was developed and used in our laboratory to identify anti-nuclear antibodies in patients sera on the basis of immunofluorescence images, and an *auc* of 0.991 was obtained with a single - well chosen - index [[36], [37].
ii. estimate the information that might be obtained by determining an unknown parameter in a given patient, e.g. by comparing results shown on *Tables 2, 4* and *6*.
iii. More generally, chose a minimal set of biological parameters that should be measured in a patient with suspected SLE or MTCD.

## 5 Conclusion

The main purpose of this report was to explore the feasibility of using currently available machine learning algorithms to address an actual - and fairly complex - diagnosis problem. The following two points may be emphasized:

i. while a proper use of validated tools was found to allow significant, although limited, classification efficiency, it must be emphasized that a much more extensive data set would be required to improve the classifiers and validate conclusions.
ii. An important conclusion might be that the use of quantitative tools might provide new guidelines about medical diagnosis. Indeed, the fairly negative conclusions of unsupervised clustering, as described in section 3.6, may suggest that it may not be a good idea to look for efficient definition of a fuzzy entity such as MCDT, and it might be more rewarding to try and directly predict more restricted parameters such as absence of renal disease or corticosteroid sensitivity. In other words, it may not be a successful strategy to use more and more refined diagnostic tools to define an increasing and possibly excessive number of pathological categories.

## Data Availability

All data produced in the present work are contained in the manuscript.

## Authors’ contribution

NB and PB designed the study. NB coordinated data selection and immunological tests. PB performed data processing. DB, NB and PB wrote and discussed the study.

## Acknowledgment

The authors declare no no competing interest, we only benefited from institutional support and our institutions did not receive any specific funding concerning the submitted work.

